# Healthcare Resource Utilization and Costs for Patients With Eosinophilic Granulomatosis With Polyangiitis in the United States: A Retrospective Analysis of Health Insurance Claims Data

**DOI:** 10.64898/2026.04.24.26351614

**Authors:** Paul Dolin, Karina A. Keogh, Jennifer Rowell, Chris Edmonds, Danuta Kielar, Juliana Meyers, Elizabeth Esterberg, Tram Nham, Stephanie Y. Chen

**Affiliations:** BioPharmaceuticals Medical, AstraZeneca, Cambridge, UK; Division of Pulmonary and Critical Care Medicine, Mayo Clinic, Rochester, MN, USA; Market Access and Pricing, AstraZeneca, Cambridge, UK; Market Access and Pricing, AstraZeneca, Gaithersburg, MD, USA; RTI Health Solutions, Research Triangle Park, NC, USA; BioPharmaceuticals Medical, AstraZeneca, Gaithersburg, MD, USA

**Keywords:** Eosinophilic granulomatosis with polyangiitis, Healthcare costs, Healthcare resources, Insurance claims analysis, Real-world evidence, Severe uncontrolled asthma

## Abstract

**Purpose:** We evaluated healthcare resource utilization (HCRU) and costs in patients with eosinophilic granulomatosis with polyangiitis (EGPA).

**Methods:** Patients with newly diagnosed EGPA (2017–2021), ≥12 months’ pre-diagnosis health plan enrollment, and ≥1 inpatient or ≥2 outpatient claims with an EGPA diagnosis were included. Follow-up was from EGPA diagnosis until disenrollment or database end. HCRU and health insurer payment costs during follow-up were compared with those for matched cohorts of general insured patients without EGPA (comparison A) and without EGPA but with severe uncontrolled asthma (SUA; comparison B).

**Results:** In comparison A, all-cause HCRU was higher in the EGPA cohort (n = 213) versus matched patients (n = 779) for all clinical encounters/pharmacy claim types; annualized, mean total all-cause costs were 16-fold higher ($117,563/patient) versus matched patients ($7,520/patient). In comparison B, all-cause HCRU was higher for the EGPA cohort (n = 182) versus the matched SUA cohort (n = 640) for all clinical encounters/pharmacy claim types, with 5-fold higher mean total all-cause costs ($118,127/patient vs $22,286/patient). In both EGPA cohorts, HCRU and associated costs increased between the baseline and follow-up periods.

**Conclusions:** These findings highlight the need for more effective treatments to reduce the clinical and economic burden of EGPA.

## Introduction

Eosinophilic granulomatosis with polyangiitis (EGPA) is a rare, small- to medium-sized vessel vasculitis characterized by polysystemic eosinophil-rich granulomatous inflammation, eosinophilia, and asthma.^1,2^ EGPA can manifest with a relapsing and remitting course in multiple tissues^3–6^, leading to multi-organ damage and disability.^1,7–10^ The disease is currently poorly understood, with limited research aimed at optimizing the comprehension of its intricate pathogenesis and effective management strategies.^11^

Treatment of EGPA – usually with oral glucocorticoids (OGCs) alone or in combination with immunosuppressive therapies – aims to induce remission, and reduce disease relapses, thereby limiting long-term organ damage progression.^1,12^

However, reliance on OGC-based treatments, often at high doses and/or for prolonged periods, puts patients at high risk of adverse effects, such as diabetes and osteoporosis.^14^

More recently, the management of EGPA has evolved to include monoclonal antibody therapies targeting interleukin-5 or its receptor (anti-IL-5/R), such as mepolizumab^15–17^ and benralizumab,^15^ both of which are approved for patients with EGPA.^12,18^ Rituximab is also recommended off-label for remission induction and maintenance in severe EGPA.^12,18^

With conventional EGPA therapies, remission rates are relatively low, and patients may experience relapses of vasculitis or asthma symptoms when attempting to taper off OGCs.^7,19 19^ The high rate of relapse adds to the burden of the disease, as well as being associated with increased healthcare resource utilization (HCRU) and costs due to excessive symptoms, associated comorbidities, and treatment requirements.^20,21^ Although EGPA is rare, with an estimated prevalence of 15.27 cases per million globally^21^ and from 3.2–30.7 cases per million in the US,^22^ the high clinical burden associated with the disease is notable,^20–24^ and limited data exist concerning the overall (clinical and economic) burden of EGPA. This is largely explained by the historical absence of relevant diagnostic codes to identify patients with EGPA from electronic health records. Only in 2015 was a unique diagnostic code established in the US within the *International Classification of Diseases, Tenth Revision, Clinical Modification* (ICD-10-CM).^20^ Therefore, further real-world data on patient characteristics, HCRU, and health insurer payment costs in patients with EGPA are needed to evaluate the full impact of the disease.

The aim of this study was to quantify all-cause HCRU and payor healthcare costs of patients in the US with EGPA and to evaluate the costs of HCRU for patients with EGPA compared with those without EGPA. Additionally, due to the potential overlap in disease characteristics and treatment between EGPA and asthma – since most patients with EGPA also have asthma and both conditions are treated with type 2 biologics – we included a second comparator group consisting of patients with severe uncontrolled asthma (SUA).^25^

## Methods

### Study Design

This study was a retrospective analysis of US administrative health insurance claims data from the Merative^TM^ MarketScan® Commercial Claims and Encounters database and the Medicare Supplemental and Coordination of Benefits (Medicare) database. The study forms part of the CONSTELLATION real-world evidence program in rare, eosinophil-associated diseases. An EGPA cohort was defined among patients with EGPA using the ICD-10-CM code M30.1 (see **Supplementary Methods**) newly diagnosed between January 1, 2017, and June 30, 2021, who were followed from the date of their first claim with a diagnosis of EGPA (index date [ID]) to the end of the database (June 30, 2021), or until disenrollment from their health plan if this occurred earlier (the “Full Incident EGPA Cohort”) (**Supplementary Figure 1**). Furthermore, patients in the Full Incident EGPA Cohort were required to have ≥12 months of continuous health plan enrolment before ID. The 12-month period prior to ID defined the baseline period. Patients with the EGPA code prior to January 1, 2017, or a first diagnosis of microscopic polyarteritis nodosa, giant cell arteritis, Takayasu arteritis, or hypereosinophilic syndrome after the last observed EGPA diagnosis were excluded.

From the Full Incident EGPA Cohort, two matched cohorts were formed based on the availability of matched control patients: Matched EGPA Cohort A (matched to the General Insured Cohort) and Matched EGPA Cohort B (matched to the SUA Cohort); both Matched EGPA Cohorts A and B included patients with and without SUA. A direct covariate matching method was used to generate a Matched General Insured Cohort by random selection of up to four persons who had no diagnosis of EGPA at any time in the claims database and otherwise matched a patient in the EGPA cohort for selected criteria (ie, sex, payor type, plan type, geographic region, age at ID [±2 years], and duration of continuous plan membership before and after ID). Each matched, general insured patient was assigned a pseudo-ID, which was the same as the ID of the matched EGPA patient. The Matched SUA Cohort was generated by random selection of up to four persons in the claims database who met the SUA definition criteria (see **Supplementary Methods**), had no diagnosis of EGPA at any time, and had an ID defined based on meeting SUA criteria on or after January 1, 2017. Matching was performed using the calendar year of the ID in addition to the criteria that had been used the Matched General Insured Cohort. The non-EGPA patient was required to have a duration of membership after the ID (follow-up period) equal to or longer than that of the EGPA patient (if the non-EGPA patient had a longer membership duration, this was censored to ensure it was exactly of equal duration after the ID to that of the EGPA patient).

### Ethics

RTI International’s Institutional Review Board evaluated this study and classified it as not meeting the regulatory requirements for research with human subjects, as data were pre-existing and de-identified. Data were anonymized and compliant with the patient requirements of the Health Insurance Portability and Accountability Act (HIPAA).^26^

### Study Outcomes

Patient demographics and characteristics were reported, including for the comorbidities listed in **Supplementary Table 1**. Study outcomes included all-cause HCRU and health insurer payment costs, analyzed overall and by service setting, for all study cohorts. Service settings included urgent care centers, which deliver urgent ambulatory care in a dedicated medical facility outside of a traditional emergency department. Mean length of stay per hospital inpatient visit was defined as the average number of inpatient days per hospitalization, calculated as the total number of inpatient days divided by the total number of inpatient admissions. Pharmacy claims included pharmacy billing for dispensed drugs; billing for drugs and their administration in hospital outpatient services were included in hospital outpatient costs. All-cause outcomes included all claims regardless of the diagnoses, procedures, or medications.

### Definitions

#### Asthma Comorbidity Definitions

A patient was defined as having persistent asthma if they met any of the following criteria: 1) ≥1 claim for an emergency department visit with a diagnosis of asthma (ICD-10-CM code J45.xx) in any position on the claim; 2) ≥1 claim for hospitalization with asthma as the primary diagnosis on the claim; 3) ≥4 outpatient claims on different dates (≥7 days apart) with an asthma diagnosis in any position on the claim and ≥2 pharmacy claims for asthma controller medications (ie, inhaled corticosteroid [ICS], ICS in combination with a long-acting β_2_ agonist [LABA], leukotriene receptor antagonist, and theophylline) in 1 year; or 4) ≥4 pharmacy claims for any asthma medication and ≥1 outpatient claim with an asthma diagnosis in any position within 1 year, similar to the criteria previously described.

A patient was defined as having severe asthma if they had persistent asthma (as defined previously) plus either: 1) receipt of ≥1 claim with a medium to high dose of ICS as defined in Global Initiative for Asthma guidelines ^27^ in combination with a LABA, long-acting muscarinic antagonist, or leukotriene receptor antagonist; or 2) ≥180 cumulative days’ supply of OGCs (gaps present between filled prescriptions that totaled ≥180 days were allowed).

A patient was defined as having SUA if they had severe asthma (as defined prior) plus any of the following: 1) ≥1 claim for hospitalization with asthma as the primary diagnosis on the claim; 2) ≥2 outpatient visits (including emergency department or urgent care) for asthma, followed by treatment with systemic glucocorticoids within 1 week (one dose of intravenous glucocorticoids or OGCs for ≥3 days); or 3) ≥4 claims for a short-acting β2 agonist within any 12-month period.

#### Disease Status Definitions

Remission was defined as no new BVAS event (as defined using diagnostic codes shown in **Supplementary Table 2**) and OGC ≤4 mg/day ^15,28^ with none of the following: OGC dose increase; use of intravenous methylprednisolone; initiation or switching of immunosuppressants or biologics (except for switching from cyclophosphamide to another immunosuppressant); or use of intravenous immunoglobulins or plasma exchange.

Major active disease was defined as the occurrence of any BVAS event, asthma exacerbation, or nasal polyp and use of cyclophosphamide or non-prophylactic use of intravenous glucocorticoids in the prior 3 months; or ≥1 major BVAS event, or ≥2 minor events in ≥2 organ systems in the prior 3 months and OGC >7.5mg/day at diagnosis. Patients who were hospitalized at ID were included with patients with major active disease (as defined previously) for analysis.

Non-major active disease was defined as every status not classified as remission, major active disease, or hospitalization.

### Statistical Analyses

Patient demographics, clinical characteristics, and HCRU outcomes are presented using descriptive analyses. The statistical significance of differences in study measures and outcomes between the EGPA and matched cohorts was measured as follows:

- Co-morbidities at baseline were compared between the EGPA and matched non-EGPA cohorts using prevalence ratios, with 95% confidence intervals (CIs) and *P*-values (estimated using Zou’s modified Poisson regression).^29^
- Annualized, all-cause HCRU during follow-up was compared between the EGPA and matched non-EGPA cohorts using rate ratios with 95% CIs and *P*-values (estimated using Poisson regression).^30^
- Annualized, all-cause healthcare costs during follow-up were compared between the EGPA and matched non-EGPA cohorts using both a cost ratio with 95% CIs and a cost difference with 95% CIs and *P*-values (estimated using Satterthwaite t-tests to account for unequal variances).

HCRU and associated health insurer payment costs (not including deductibles, co-pay, or co-insurance) were calculated per patient per year for the 12-month baseline period and for the total follow-up period (as an annualized value). Per patient per year costs were calculated at the patient level then averaged across patients with no further weighting applied. Healthcare costs were adjusted to 2021 US dollars using the medical care component of the US Consumer Price Index. All analyses were conducted using SAS® statistical software (version 9.4 or later; SAS Institute, Cary, NC, USA).

## Results

### Baseline Demographics and Characteristics

In total, there were 236 patients with incident EGPA who were newly diagnosed from 2017–2021 in the Full Incident EGPA cohort, of whom 213 (90.3%) comprised Matched EGPA Cohort A and 182 (77.1%) comprised Matched EGPA Cohort B (**Supplementary Figure 2**).

#### Matched EGPA Cohort A and the Matched General Insured Cohort

Both Matched EGPA Cohort A (n = 213) and the Matched General Insured Cohort (n = 779) had a similar demographic profile as the Full Incident EGPA Cohort (**Supplementary Table 3**). Most patients in the Matched EGPA Cohort A and the Matched General Insured Cohort were female (59%) and had commercial insurance (90%); the mean age at ID was similar between cohorts (49.6 and 50.6 years in Matched EGPA Cohort A and the Matched General Insured Cohort, respectively). The mean follow-up duration was 21 months in each matched cohort.

Matched EGPA Cohort A had a higher prevalence of comorbidities in the 12 months prior to the ID than the Matched General Insured Cohort. Of the 28 co-morbidities examined with prevalence ≥1% in either cohort, there were significant differences between the Matched EGPA and Matched General Insured cohorts in the prevalence of asthma (including SUA), throat or chest pain, gastroesophageal reflux disease, nasal polyps, vitamin D deficiency, chronic obstructive pulmonary disease, arrhythmia, atopic dermatitis/eczema, ischemic heart disease, pulmonary eosinophilia, interstitial pulmonary disease, deep vein thromboembolism, primary (essential) thrombocythemia, and hyperthyroidism. The most prevalent comorbidities in Matched EGPA Cohort A were asthma related; persistent and severe asthma were approximately 26- and 57-fold more prevalent, respectively, in Matched EGPA Cohort A than in the Matched General Insured Cohort (**Supplementary Table 4**).

#### Matched EGPA Cohort B and the Matched SUA Cohort

Both Matched EGPA Cohort B (n = 182) and the Matched SUA Cohort (n = 640) had baseline characteristics similar to those of the Full Incident EGPA Cohort (**Supplementary Table 5**). The majority of patients in the matched cohorts were female (∼60%) and had commercial insurance (89%); the mean ages at ID were well matched (51.2 and 51.7 years in Matched EGPA Cohort B and the Matched SUA Cohort, respectively). The mean follow-up duration was 19.9 months in Matched EGPA Cohort B and 18.8 months in the Matched SUA Cohort.

In the 12 months before ID, 27 of the co-morbidities examined (in addition to the co-morbidity of asthma that was an inclusion criterion for the cohort) occurred in ≥1% of patients in either Matched EGPA Cohort B or the Matched SUA Cohort. The majority of co-morbidities showed comparable prevalence in both cohorts; the most common was dyslipidemia (43.4% and 48.9% in the EGPA and SUA cohorts, respectively). A significant difference was seen in the prevalence of nasal polyps, obesity, arrhythmia, pulmonary eosinophilia, and interstitial pulmonary disease. A majority of, but not all, patients in the EGPA cohort had persistent (70.3%) or severe (64.3%) asthma (**Supplementary Table 6**).

### Healthcare Resource Utilization

During the follow-up period, in the Matched EGPA Cohort A versus the Matched General Insured Cohort (**Figure 1A**), annualized, all-cause HCRU were higher for all types of clinical encounters, and in Matched EGPA Cohort B versus the Matched SUA Cohort were higher for most of the clinical encounters (hospital inpatient visits, hospital outpatient visits, ambulatory surgical center visits, physician office visits, home visits, telemedicine, and other outpatient visits and ancillary care (**Figure 1B**).

**Figure 1.**
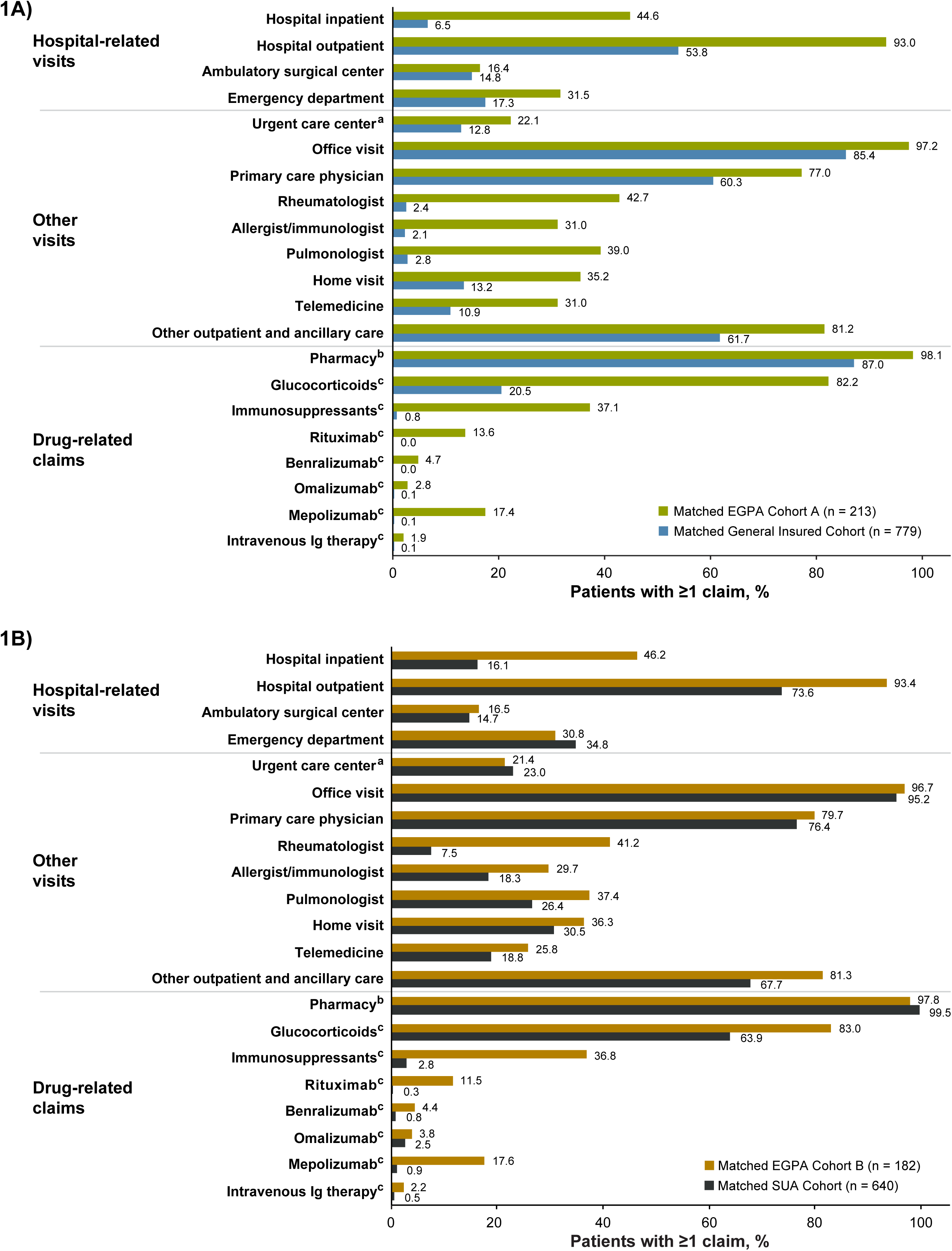
Annualized, all-cause healthcare resource utilization (% patients with ≥1 claim) during the total follow-up period **(A)** in Matched EGPA Cohort A and the Matched General Insured Cohort, and **(B)** in Matched EGPA Cohort B and the Matched SUA cohort. **Notes:** ^a^Urgent care centers deliver urgent ambulatory care in a dedicated medical facility outside of a traditional emergency department. ^b^Pharmacy costs cover pharmacy billing for dispensed drugs. ^c^Includes pharmacy claims and medical claims in hospital inpatient and outpatient settings. **Abbreviations:** EGPA, eosinophilic granulomatosis with polyangiitis; Ig, immunoglobulin; SUA, severe uncontrolled asthma.

#### Matched EGPA Cohort A and the Matched General Insured Cohort

With respect to the proportion of patients with visits to healthcare professionals, differences between the cohorts were most marked for hospital inpatient visits (44.6% vs 6.5%); hospital outpatient visits (93.0% vs 53.8%); emergency department visits (31.5% vs 17.3%); urgent care center visits (22.1% vs 12.8%); office visits to a rheumatologist (42.7% vs 2.4%), allergist/immunologist (31.0% vs 2.1%), or pulmonologist (39.0% vs 2.8%); home visits (35.2% vs 13.2%); and telemedicine appointments (31.0% vs 10.9%). HCRU (the proportion of patients with a visit) to ambulatory surgical centers was similar in both cohorts. Regarding drug-related claims, HCRU for each of glucocorticoids, immunosuppressants, biologics, and intravenous immunoglobulins was notably higher in EGPA Cohort A than in the Matched General Insured Cohort.

The annualized mean number of all-cause claims/patient/year in Matched EGPA Cohort A was higher in the follow-up period after their diagnosis versus the pre-diagnosis baseline period for inpatient visits, hospital outpatient visits, office visits, home visits, and combined other outpatient visits and ancillary care claims (**Figure 2A**). The mean (standard deviation [SD]) length of stay per inpatient hospital visit in Matched EGPA Cohort A was 8.2 (7.1) days before ID, compared with 16.0 (35.3) days in the follow-up period (**Figure 2B**). All-cause HCRU was generally similar between the baseline and follow-up periods in the Matched General Insured Cohort (**Figure 2A** and **2B**).

**Figure 2.**
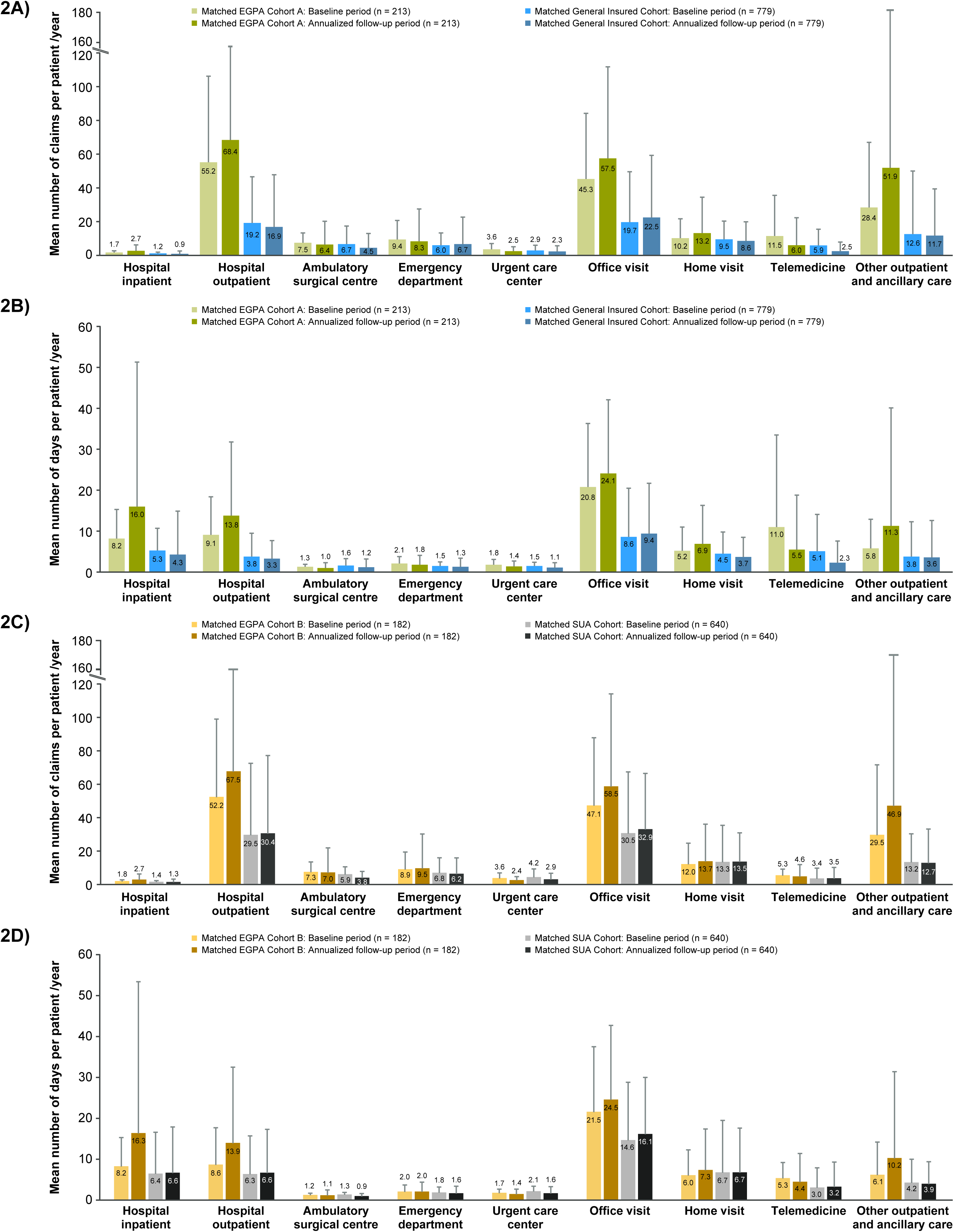
Mean all-cause (**A**) claims for visits/health care encounters in Matched EGPA Cohort A and the Matched General Insured Cohort; (**B**) days with a claim for a visit/health care encounter in Matched EGPA Cohort A and the Matched General Insured Cohort; (**C**) claims for a visit/health care encounter in Matched EGPA Cohort B and the Matched SUA Cohort; and (**D**) days with a claim for a visit/health care encounter in Matched EGPA Cohort B and the Matched SUA Cohort during the baseline period and annualized follow-up period. **Note:** Urgent care centers deliver urgent ambulatory care in a dedicated medical facility outside of a traditional emergency department. **Abbreviations:** EGPA, eosinophilic granulomatosis with polyangiitis; SUA, severe uncontrolled asthma.

#### Matched EGPA Cohort B and the Matched SUA Cohort

The all-cause mean number of claims/patient/year for hospital outpatient visits, ambulatory surgical center visits, emergency department visits, physician office visits, telemedicine, and other outpatient visits and ancillary care was higher for Matched EGPA Cohort B compared with the Matched SUA cohort at baseline and 1-year follow-up (**Figure 2C**). Patients with EGPA, compared with those with SUA, had a higher all-cause mean number of days/patient/year with a hospital inpatient visit, hospital outpatient visit, physician office visit, telemedicine, or other outpatient and ancillary care visit at baseline and 1-year follow-up (**Figure 2D**). The mean (SD) length of stay per hospital inpatient visit in the Matched EGPA Cohort B was 8.2 (7.1) days in the baseline period compared with 16.3 (37.1) days in the follow-up period. The corresponding mean length of stays in the Matched SUA Cohort were 6.4 (10.2) and 6.6 (11.3) days, respectively (**Figure 2D**). All-cause claims/patient/year were generally similar between the baseline period and the follow-up period in the Matched SUA Cohort (**Figure 2C**).

### Payor Healthcare Costs

#### Matched EGPA Cohort A and the Matched General Insured Cohort

Annualized, mean total all-cause health insurer payment costs per patient in the follow-up period were 16-fold higher in Matched EGPA Cohort A ($117,563) versus the Matched General Insured Cohort ($7,520); the main cost drivers in Matched EGPA Cohort A were inpatient ($46,410) and outpatient ($26,544) hospital visits and pharmacy claims ($25,016) (**Table 1**). Mean total costs/patient/year in Matched EGPA Cohort A more than doubled between the 12-month baseline period before their diagnosis ($43,420) and the follow-up period ($117,563). The equivalent costs for the Matched General Insured Cohort remained relatively stable ($6,435 and $7,520, respectively).

**Table 1.** Annualized All-Cause Costs/Patient/Year in Matched EGPA Cohort A and the Matched General Insured Cohort.

| Healthcare Costs, Mean (SD), \$ | Matched EGPA Cohort A<br>(n = 213) | | Matched General Insured<br>Cohort<br>(n = 779) | | Cost Ratio (95%<br>CI) for Annualized<br>Post-Index Period | P-value |
| --- | --- | --- | --- | --- | --- | --- |
|  | Baseline<br>Period | Annualized<br>Total Follow-up Period | Baseline<br>Period | Annualized<br>Total Follow-up Period |  |  |
| **Total** | **43,420 (51,551)** | **117,563**<br>**(221,547)<sup>d</sup>** | **6,435 (17,213)** | **7,520 (21,458)** | **15.6 (11.1, 21.5)** | **< .001** |
| Hospital-related visits |  |  |  |  |  |  |
| Hospital inpatient | 17,166 (34,936) | 46,410<br>(199,096)<sup>d</sup> | 1,275 (7,074) | 1,310 (11,576) | 35.4 (13.4, 102.2) | .001 |
| Hospital outpatient | 10,068 (16,680) | 26,544<br>(64,732)<sup>d</sup> | 1,496 (5,147) | 1,766 (6,502) | 15.0 (9.5, 22.7) | < .001 |
| Ambulatory surgical center | 621 (3,465) | 408 (1,779) | 320 (3,544) | 295 (2,441) | 1.4 (0.5, 3.7) | .449 |
| Emergency department | 1,157 (2,410) | 785 (2,670)<sup>d</sup> | 160 (704) | 242 (1,541) | 3.2 (1.6, 6.5) | .005 |
| Other visits |  |  |  |  |  |  |
| Urgent care center<sup>a</sup> | 51 (154) | 42 (130)<sup>d</sup> | 12 (55) | 16 (73) | 2.7 (1.5, 4.5) | .005 |
| Office | 4,646 (7,805) | 13,127<br>(29,593)<sup>d</sup> | 902 (2,665) | 1,222 (4,642) | 10.7 (7.0, 16.2) | < .001 |
| Home | 376 (1,702) | 1,794 (12,008)<sup>d</sup> | 59 (522) | 60 (358) | 29.9 (2.9, 69.9) | .036 |
| Telemedicine | 67 (431) | 240 (949)<sup>d</sup> | 10 (143) | 23 (223) | 10.2 (4.1, 33.1) | .001 |
| Other outpatient visits and<br>ancillary care | 1,112 (3,011) | 3,198 (13,436)<sup>d</sup> | 577 (5,688) | 283 (1,411) | 11.3 (4.7, 21.1) | .002 |
| Drug-related costs |  |  |  |  |  |  |
| Pharmacy claims<sup>b</sup> | 8,156 (15,185) | 25,016<br>(36,361)<sup>d</sup> | 1,624 (6,825) | 2,305 (12,802) | 10.9 (7.3, 18.3) | < .001 |
| Glucocorticoids | 16 (27) | 275 (2,829) | 1 (3) | 0 (2) | 688.7<br>(-275.0, 1,983.8) | .158 |
| Immunosuppressive therapies | 53 (366) | 380 (1,529)<sup>d</sup> | 7 (196) | 8 (206) | 50.6 (NE, NE) | < .001 |
| Biologics | 809 (5,418) | 258 (2,302) | 30 (846) | 9 (239) | 30.2 (NE, NE) | .116 |
| All other | 7,278 (14,299) | 24,103<br>(35,536)<sup>d</sup> | 1,586 (6,709) | 2,288 (12,788) | 10.5 (7.1, 17.9) | < .001 |
| Medical claims for drug and<br>administration<sup>c</sup> |  |  |  |  |  |  |
| Immunosuppressive therapies | 2 (29) | 251 (1,590)<sup>d</sup> | NE | 0 (0) | NE | .022 |
| Biologics | 2,248 (9,047) | 18,430<br>(54,138)<sup>d</sup> | 12 (325) | 72 (1,491) | 255.1 (NE, NE) | < .001 |
| Intravenous Ig therapy | 152 (2,212) | 813 (9,933) | 0 (NE) | 3 (94) | 242.6 (NE, NE) | 0.236 |

**Notes:** Costs do not include payments made by patients, such as deductibles, copay, or co-insurance.
<sup>a</sup>Urgent care centers deliver urgent ambulatory care in a dedicated medical facility outside of a traditional emergency department.
<sup>b</sup>Pharmacy costs cover pharmacy billing for dispensed drugs.
<sup>c</sup>Includes inpatient and outpatient costs.
<sup>d</sup>Difference in post-index annualized costs between Matched EGPA Cohort A and the Matched General Insured Cohort was statistically significant at a threshold of $P \leq .05$ .
**Abbreviations:** CI, confidence interval; EGPA, eosinophilic granulomatosis with polyangiitis; Ig, immunoglobulin; NE, not estimable; SD, standard deviation.

#### Matched EGPA Cohort B and the Matched SUA Cohort

Annualized, mean total all-cause health insurer payment costs per patient in the follow-up period were 5-fold higher in Matched EGPA Cohort B ($118,127) versus the Matched SUA Cohort ($22,286); the main cost drivers in Matched EGPA Cohort B were inpatient ($47,980) and outpatient ($27,714) hospital visits and pharmacy claims ($24,753) (**Table 2**). The mean total costs/patient/year in Matched EGPA Cohort B more than doubled between the 12-month baseline period ($41,837) and the follow-up period ($118,127). The equivalent costs for the Matched SUA Cohort remained relatively stable, with only a small increase observed between the baseline and follow-up periods ($17,289 and $22,286, respectively).

**Table 2.**
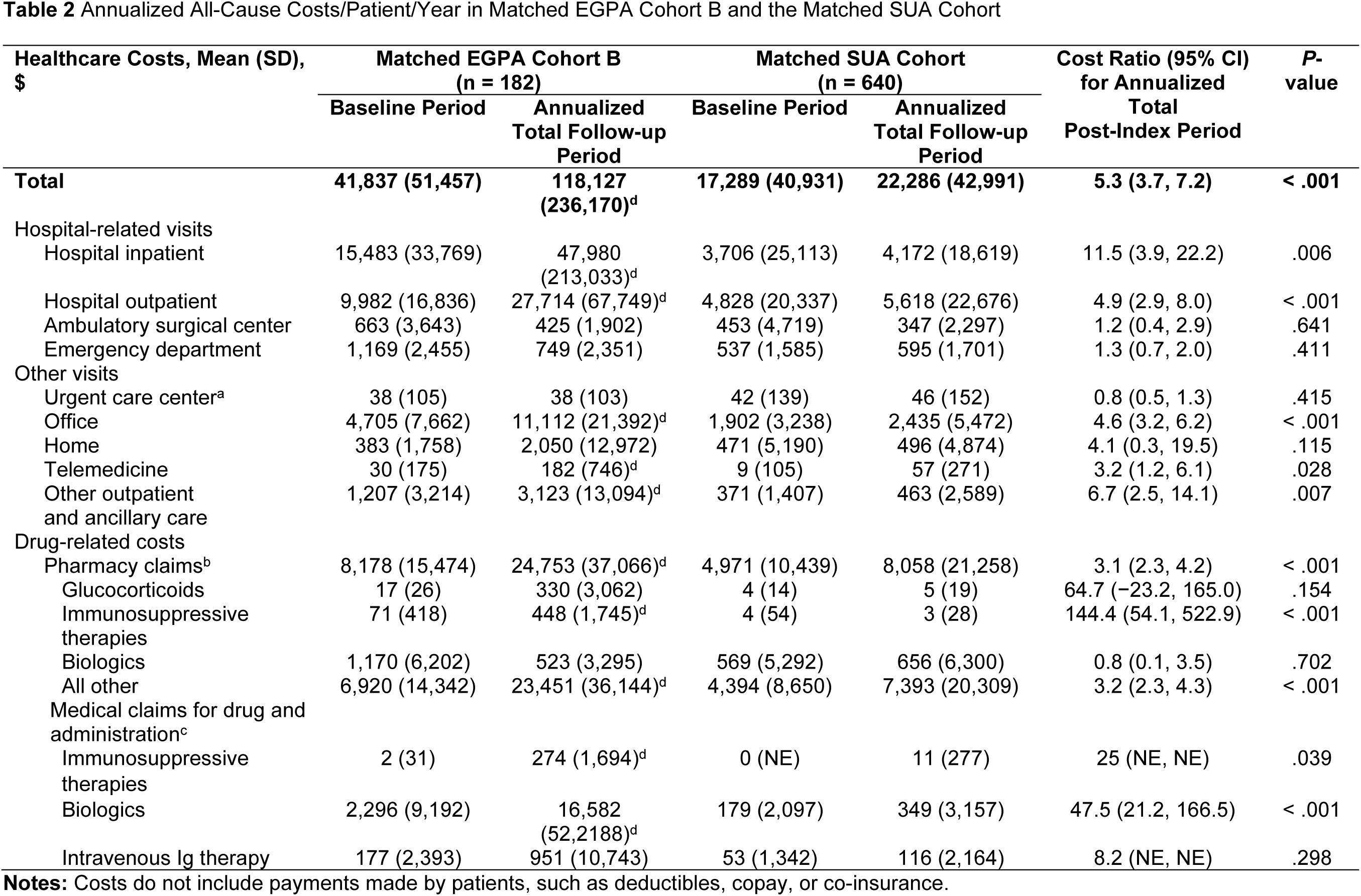
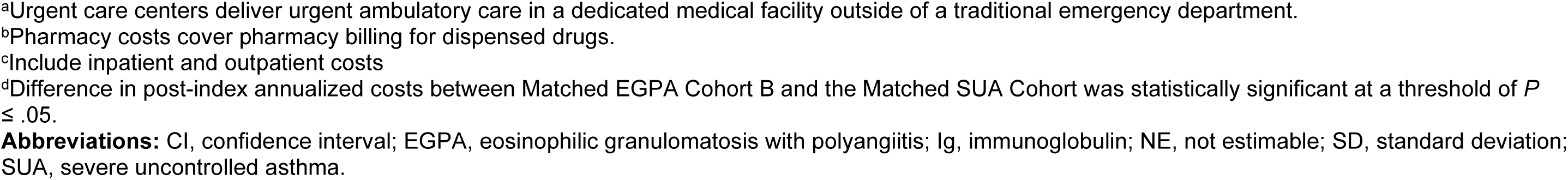
Annualized All-Cause Costs/Patient/Year in Matched EGPA Cohort B and the Matched SUA Cohort.

| Healthcare Costs, Mean (SD),<br>\$ | Matched EGPA Cohort B<br>(n = 182) | | Matched SUA Cohort<br>(n = 640) | | Cost Ratio (95% CI)<br>for Annualized<br>Total<br>Post-Index Period | P-value |
| --- | --- | --- | --- | --- | --- | --- |
|  | Baseline Period | Annualized<br>Total Follow-up<br>Period | Baseline Period | Annualized<br>Total Follow-up<br>Period |  |  |
| **Total** | **41,837 (51,457)** | **118,127**<br>**(236,170)<sup>d</sup>** | **17,289 (40,931)** | **22,286 (42,991)** | **5.3 (3.7, 7.2)** | **< .001** |
| Hospital-related visits |  |  |  |  |  |  |
| Hospital inpatient | 15,483 (33,769) | 47,980<br>(213,033)<sup>d</sup> | 3,706 (25,113) | 4,172 (18,619) | 11.5 (3.9, 22.2) | .006 |
| Hospital outpatient | 9,982 (16,836) | 27,714 (67,749)<sup>d</sup> | 4,828 (20,337) | 5,618 (22,676) | 4.9 (2.9, 8.0) | < .001 |
| Ambulatory surgical center | 663 (3,643) | 425 (1,902) | 453 (4,719) | 347 (2,297) | 1.2 (0.4, 2.9) | .641 |
| Emergency department | 1,169 (2,455) | 749 (2,351) | 537 (1,585) | 595 (1,701) | 1.3 (0.7, 2.0) | .411 |
| Other visits |  |  |  |  |  |  |
| Urgent care center<sup>a</sup> | 38 (105) | 38 (103) | 42 (139) | 46 (152) | 0.8 (0.5, 1.3) | .415 |
| Office | 4,705 (7,662) | 11,112 (21,392)<sup>d</sup> | 1,902 (3,238) | 2,435 (5,472) | 4.6 (3.2, 6.2) | < .001 |
| Home | 383 (1,758) | 2,050 (12,972) | 471 (5,190) | 496 (4,874) | 4.1 (0.3, 19.5) | .115 |
| Telemedicine | 30 (175) | 182 (746)<sup>d</sup> | 9 (105) | 57 (271) | 3.2 (1.2, 6.1) | .028 |
| Other outpatient<br>and ancillary care | 1,207 (3,214) | 3,123 (13,094)<sup>d</sup> | 371 (1,407) | 463 (2,589) | 6.7 (2.5, 14.1) | .007 |
| Drug-related costs |  |  |  |  |  |  |
| Pharmacy claims<sup>b</sup> | 8,178 (15,474) | 24,753 (37,066)<sup>d</sup> | 4,971 (10,439) | 8,058 (21,258) | 3.1 (2.3, 4.2) | < .001 |
| Glucocorticoids | 17 (26) | 330 (3,062) | 4 (14) | 5 (19) | 64.7 (-23.2, 165.0) | .154 |
| Immunosuppressive<br>therapies | 71 (418) | 448 (1,745)<sup>d</sup> | 4 (54) | 3 (28) | 144.4 (54.1, 522.9) | < .001 |
| Biologics | 1,170 (6,202) | 523 (3,295) | 569 (5,292) | 656 (6,300) | 0.8 (0.1, 3.5) | .702 |
| All other | 6,920 (14,342) | 23,451 (36,144)<sup>d</sup> | 4,394 (8,650) | 7,393 (20,309) | 3.2 (2.3, 4.3) | < .001 |
| Medical claims for drug and<br>administration<sup>c</sup> |  |  |  |  |  |  |
| Immunosuppressive<br>therapies | 2 (31) | 274 (1,694)<sup>d</sup> | 0 (NE) | 11 (277) | 25 (NE, NE) | .039 |
| Biologics | 2,296 (9,192) | 16,582<br>(52,2188)<sup>d</sup> | 179 (2,097) | 349 (3,157) | 47.5 (21.2, 166.5) | < .001 |
| Intravenous Ig therapy | 177 (2,393) | 951 (10,743) | 53 (1,342) | 116 (2,164) | 8.2 (NE, NE) | .298 |

**Notes:** Costs do not include payments made by patients, such as deductibles, copay, or co-insurance.
<sup>a</sup>Urgent care centers deliver urgent ambulatory care in a dedicated medical facility outside of a traditional emergency department.
<sup>b</sup>Pharmacy costs cover pharmacy billing for dispensed drugs.
<sup>c</sup>Include inpatient and outpatient costs
<sup>d</sup>Difference in post-index annualized costs between Matched EGPA Cohort B and the Matched SUA Cohort was statistically significant at a threshold of $P \leq .05$ .
**Abbreviations:** CI, confidence interval; EGPA, eosinophilic granulomatosis with polyangiitis; Ig, immunoglobulin; NE, not estimable; SD, standard deviation; SUA, severe uncontrolled asthma.

#### EGPA and Disease Status

Annualized all-cause costs/patient/year in Matched EGPA Cohort A were also evaluated in terms of disease status at ID (**Table 3**). For the 12-month baseline period, mean total costs were lowest among those not documented to have any evidence of disease activity during that time period (presumed remission) ($30,514) compared with those documented to have non-major active disease or major active disease at ID ($40,457 and $48,840, respectively).

**Table 3.**
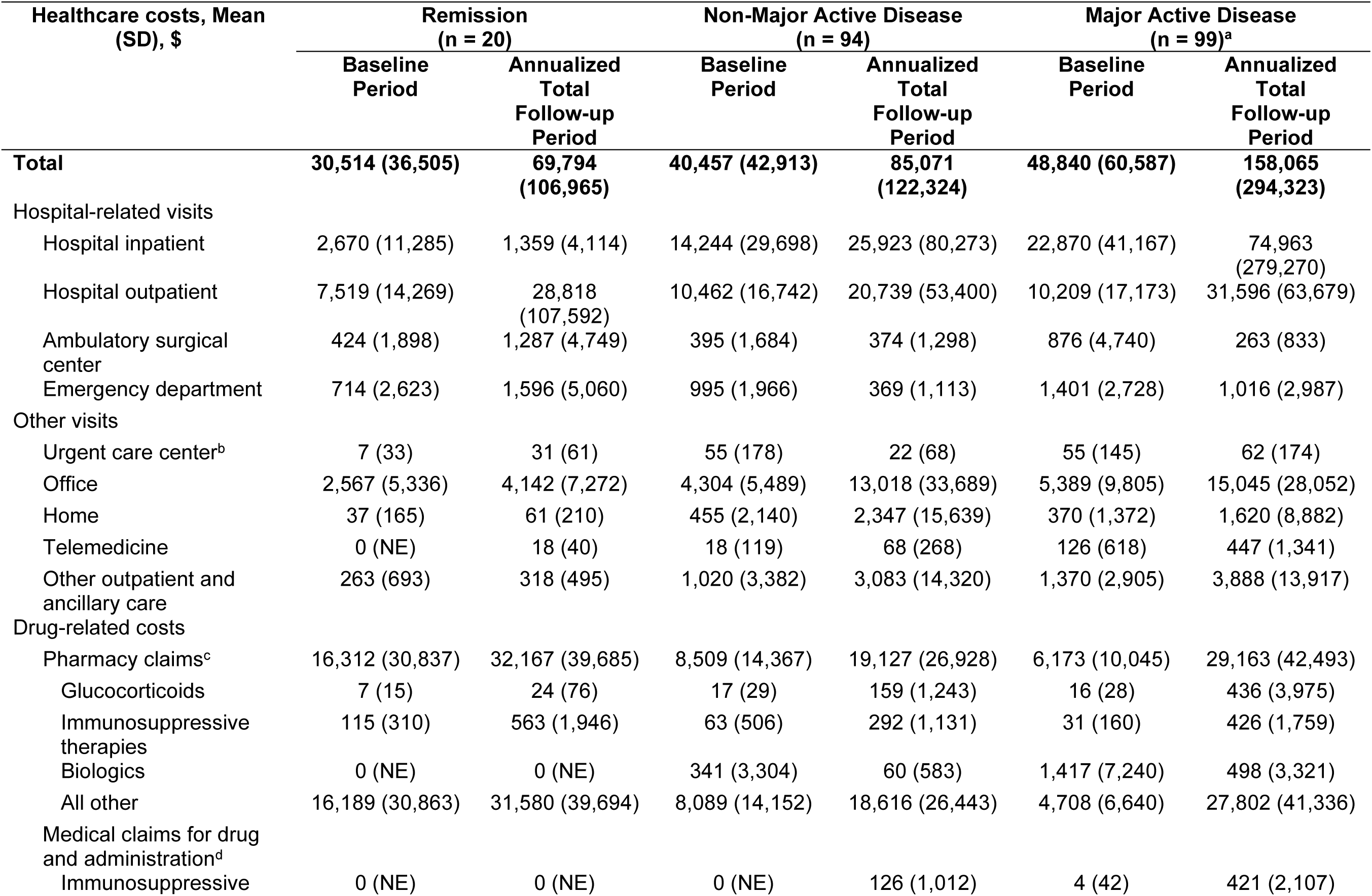
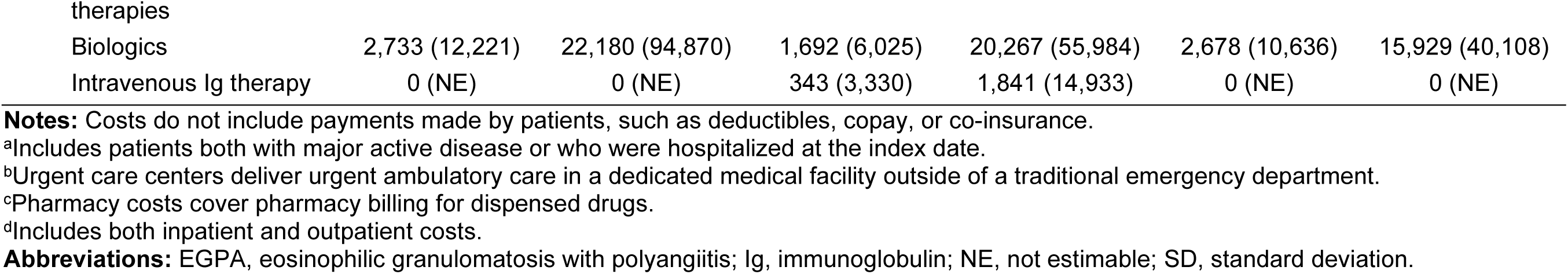
Annualized All-Cause Costs/Patient/Year by Disease Status at the Index Date in Matched EGPA Cohort A.

| Healthcare costs, Mean<br>(SD), \$ | Remission<br>(n = 20) | | Non-Major Active Disease<br>(n = 94) | | Major Active Disease<br>(n = 99)<sup>a</sup> | |
| --- | --- | --- | --- | --- | --- | --- |
|  | Baseline<br>Period | Annualized<br>Total<br>Follow-up<br>Period | Baseline<br>Period | Annualized<br>Total<br>Follow-up<br>Period | Baseline<br>Period | Annualized<br>Total<br>Follow-up<br>Period |
| **Total** | **30,514 (36,505)** | **69,794**<br>**(106,965)** | **40,457 (42,913)** | **85,071**<br>**(122,324)** | **48,840 (60,587)** | **158,065**<br>**(294,323)** |
| Hospital-related visits |  |  |  |  |  |  |
| Hospital inpatient | 2,670 (11,285) | 1,359 (4,114) | 14,244 (29,698) | 25,923 (80,273) | 22,870 (41,167) | 74,963<br>(279,270) |
| Hospital outpatient | 7,519 (14,269) | 28,818<br>(107,592) | 10,462 (16,742) | 20,739 (53,400) | 10,209 (17,173) | 31,596 (63,679) |
| Ambulatory surgical<br>center | 424 (1,898) | 1,287 (4,749) | 395 (1,684) | 374 (1,298) | 876 (4,740) | 263 (833) |
| Emergency department | 714 (2,623) | 1,596 (5,060) | 995 (1,966) | 369 (1,113) | 1,401 (2,728) | 1,016 (2,987) |
| Other visits |  |  |  |  |  |  |
| Urgent care center<sup>b</sup> | 7 (33) | 31 (61) | 55 (178) | 22 (68) | 55 (145) | 62 (174) |
| Office | 2,567 (5,336) | 4,142 (7,272) | 4,304 (5,489) | 13,018 (33,689) | 5,389 (9,805) | 15,045 (28,052) |
| Home | 37 (165) | 61 (210) | 455 (2,140) | 2,347 (15,639) | 370 (1,372) | 1,620 (8,882) |
| Telemedicine | 0 (NE) | 18 (40) | 18 (119) | 68 (268) | 126 (618) | 447 (1,341) |
| Other outpatient and<br>ancillary care | 263 (693) | 318 (495) | 1,020 (3,382) | 3,083 (14,320) | 1,370 (2,905) | 3,888 (13,917) |
| Drug-related costs |  |  |  |  |  |  |
| Pharmacy claims<sup>c</sup> | 16,312 (30,837) | 32,167 (39,685) | 8,509 (14,367) | 19,127 (26,928) | 6,173 (10,045) | 29,163 (42,493) |
| Glucocorticoids | 7 (15) | 24 (76) | 17 (29) | 159 (1,243) | 16 (28) | 436 (3,975) |
| Immunosuppressive<br>therapies | 115 (310) | 563 (1,946) | 63 (506) | 292 (1,131) | 31 (160) | 426 (1,759) |
| Biologics | 0 (NE) | 0 (NE) | 341 (3,304) | 60 (583) | 1,417 (7,240) | 498 (3,321) |
| All other | 16,189 (30,863) | 31,580 (39,694) | 8,089 (14,152) | 18,616 (26,443) | 4,708 (6,640) | 27,802 (41,336) |
| Medical claims for drug<br>and administration<sup>d</sup> |  |  |  |  |  |  |
| Immunosuppressive | 0 (NE) | 0 (NE) | 0 (NE) | 126 (1,012) | 4 (42) | 421 (2,107) |
| Biologics | 2,733 (12,221) | 22,180 (94,870) | 1,692 (6,025) | 20,267 (55,984) | 2,678 (10,636) | 15,929 (40,108) |
| Intravenous Ig therapy | 0 (NE) | 0 (NE) | 343 (3,330) | 1,841 (14,933) | 0 (NE) | 0 (NE) |

**Notes:** Costs do not include payments made by patients, such as deductibles, copay, or co-insurance.
<sup>a</sup>Includes patients both with major active disease or who were hospitalized at the index date.
<sup>b</sup>Urgent care centers deliver urgent ambulatory care in a dedicated medical facility outside of a traditional emergency department.
<sup>c</sup>Pharmacy costs cover pharmacy billing for dispensed drugs.
<sup>d</sup>Includes both inpatient and outpatient costs.
**Abbreviations:** EGPA, eosinophilic granulomatosis with polyangiitis; Ig, immunoglobulin; NE, not estimable; SD, standard deviation.

Compared with the baseline period, mean total costs in the follow-up period approximately doubled for those with no documented disease activity (presumed remission) and non-major active disease, and they approximately tripled for those with major active disease. Across all subgroups, the major cost drivers during the follow-up period were hospital outpatient visits, pharmacy claims, and office visits; for the non-major active disease and major active disease subgroups, inpatient hospitalization was the largest driver ($25,923 and $74,963, respectively).

## Discussion

There is a scarcity of data concerning the real-world clinical and economic burden of EGPA. In addressing this crucial data gap, the current retrospective analysis aimed to provide a comprehensive description of the healthcare and economic burden associated with EGPA in the US. Our objective was to utilize extensive data acquired from a US claims database to identify and characterize well-defined cohorts of patients diagnosed with EGPA and compare them with cohorts of patients without EGPA, or without EGPA but with SUA.

During the 12 months leading up to ID (baseline period), Matched EGPA Cohort A exhibited a higher prevalence of comorbidities compared with the Matched General Insured Cohort. It is notable that the reported percentage of the most prevalent comorbidity in the EGPA cohort (ie, persistent asthma, reported for 70.4% of patients) may actually be an underestimation of the real value, as an individual’s asthma might not have been included in the dataset if it had been diagnosed outside the study period or if their condition was well managed and did not require visits to a healthcare professional.

Considering the history of comorbidities in Matched EGPA Cohort B versus the Matched SUA Cohort, it is important to note that while some of these comorbidities were linked to EGPA, others might have been attributed to the long-term use of OGCs. Overall, the significant prevalence of respiratory and systemic comorbidities among EGPA patients and the associated risk emphasize the need for a proactive response to treatment needs.

The retrospective analysis of insurance claims data showed that the annualized, mean all-cause costs/patient/year were much higher for patients with EGPA than for demographically matched patients in the general insured population or those with SUA ($117,563 vs $7,520 and $118,127 vs $22,286, respectively); the costs identified in this study were of similar magnitude to those identified in prior studies.^24,31^ These increased costs for patients with EGPA, especially for those with active disease at ID, reflect a high disease burden on patients and healthcare systems alike, and they were observed across most types of healthcare visits and clinical encounters. Compared with the Matched General Insured Cohort, more healthcare visits and longer hospital stays were observed in Matched EGPA Cohort A following diagnosis. The increase in healthcare encounters may be attributed to the necessity of specialized consultations post diagnosis, while prolonged hospitalization may be a consequence of involving multiple specialists owing to the multi-organ involvement characteristic of EGPA. Nevertheless, the higher follow-up costs that were observed in patients with major active disease at baseline, compared with those with remission and non-major active disease, demonstrate the importance of preventing major events. Overall, such findings are consistent with those of a small number of previous studies conducted in the US and Europe that reported on the high burden of EGPA.^17,20–22^

Some limitations of this study relate to the nature of the analysis, which was a secondary retrospective analysis of insurance data collected for billing purposes. The EGPA cohort was defined solely based on the ICD-10 diagnosis code, without considering treatment history, which may have led to the inclusion of misdiagnosed individuals. The requirement for one inpatient EGPA diagnosis or two outpatient records of EGPA diagnosis, spaced more than three months apart, alongside the exclusion of patients diagnosed with microscopic polyarteritis nodosa, giant cell arteritis, Takayasu arteritis, or hypereosinophilic syndrome after the last observed EGPA diagnosis, aimed to reduce this risk. Nevertheless, some patients without EGPA, such as those with other types of ANCA-associated vasculitis, may still have been included. Furthermore, there may have been under coding or misclassification due to the use of diagnosis codes as a proxy for BVAS measures. We assumed non-informative censoring; however, if patients lost to follow-up differed substantially from those remaining under observation – particularly if censoring was associated with greater disease severity – cost and HCRU estimates may have been biased. Although EGPA in the era of modern treatment is not associated with high mortality, with five-year survival rates of approximately 90%, loss to follow-up may still disproportionately affect patients with more severe or complex disease who incur higher healthcare utilization. Consequently, the economic burden reported here may have been underestimated. Additionally, services paid out-of-pocket, uninsured care, or treatments received outside the covered network are not included, which could lead to underestimation of HCRU and costs. Variability in medical coding practices, including up-coding or under-coding, can introduce inaccuracies in HCRU and cost estimation. Additionally, patients may switch insurance providers or plans, which leads to gaps in data continuity and loss to follow-up. Since the data collection period, the use of biologic anti-IL-5/R therapy for severe asthma has increased, likely impacting the current costs for individuals with severe asthma. Another limitation is related to the relapsing, remitting nature of the disease, which made it difficult to definitively ascertain the date of EGPA onset and any late relapses, long-term cumulative costs and chronic complications of OGC use. Furthermore, the disease statuses of remission, non-major active disease, and relapse in the claims data were based on proxies and not based on clinical results or measurements. The use of OGCs as a proxy for relapses has some limitations. While claims for OGCs are usually an indicator of relapse as they are commonly used to manage such events,^32^ they may also be used for other conditions, which could affect the results. Additionally, adherence to prescribed OGCs was assumed, as patient compliance could not be verified. Finally, a decline in patient numbers over the follow-up period was observed, particularly in Years 3 and 4.

## Conclusions

The findings of this analysis add to the limited existing data on the clinical and economic burden of EGPA and highlight the need for more effective, targeted treatments to induce and maintain remission and reduce clinical encounters. The observed increase in costs following diagnosis is likely multifactorial, reflecting not only disease severity and progression but also diagnostic evaluation, continued care and management, and the initiation of advanced therapies.

## Abbreviations

ACR: American College of Rheumatology
anti-IL-5/R: interleukin-5 or its receptor
BVAS: Birmingham Vasculitis Activity Score
CI: confidence interval
EGPA: eosinophilic granulomatosis with polyangiitis
EULAR: European Alliance of Associations for Rheumatology
HCRU: healthcare resource utilization
HIPAA: Health Insurance Portability and Accountability Act
ICD-10-CM: *International Classification of Diseases, Tenth Revision, Clinical Modification*
ICS: inhaled corticosteroid
ID: index date
Ig: immunoglobulin
LABA: long-acting β_2_ agonist
NE: not estimable
OGC: oral glucocorticoid
SD: standard deviation
SUA: severe uncontrolled asthma
US: United States

## Acknowledgments

The authors would like to thank Anat Shavit (AstraZeneca, Cambridge, UK) for providing input on the study design and data interpretation and Bo Ding (AstraZeneca, Gothenburg, Sweden) for providing input on data interpretation. Graziella Greco and Elizabeth Strickland of Springer Health+, Springer Healthcare Ltd, UK, provided medical writing support, which was funded by AstraZeneca in accordance with Good Publication Practice 2022 guidelines.

## Disclosure

PD, JR, CE, DK, and SYC are or were employees or contractors of AstraZeneca at the time of study conduct and may own stock/stock options. DK is currently an employee of Viatris.

KAK is a site principal investigator for an AstraZeneca pharmaceutical trial in asthma.

JM, EE, and TN are employees of RTI Health Solutions, which received funding from AstraZeneca to conduct the study.

## Ethical Approval

This study was classified as not meeting the regulatory requirements for research with human subjects, as data were pre-existing and de-identified. Data were anonymized and compliant with the HIPAA requirements.

## Data Availability

The data that support the findings of this study are available from Merative^TM^, but restrictions apply to the availability of these data, which were used under license for the current study and so are not publicly available.

## Author Contributions

All authors made a significant contribution to the work reported, whether that is in the conception, study design, execution, acquisition of data, analysis and interpretation, or in all these areas; took part in drafting, revising or critically reviewing the article; gave final approval of the version to be published; have agreed on the journal to which the article has been submitted; and agree to be accountable for all aspects of the work.

## References

1. Fijolek J, Radzikowska E. Eosinophilic granulomatosis with polyangiitis: advances in pathogenesis, diagnosis, and treatment. Front Med (Lausanne). 2023;10:1145257. doi: 10.3389/fmed.2023.1145257.

2. Furuta S, Iwamoto T, Nakajima H. Update on eosinophilic granulomatosis with polyangiitis. Allergol Int. 2019;68(4):430–436. doi: 10.1016/j.alit.2019.06.004.

3. Caminati M, Fassio A, Alberici F, et al. Eosinophilic granulomatosis with polyangiitis onset in severe asthma patients on monoclonal antibodies targeting type 2 inflammation: report from the European EGPA study group. Allergy. 2024;79(2):516–519. doi: 10.1111/all.15934.

4. Hagemann J, Laudien M, Becker S, et al. EGPA: eosinophilic granulomatosis with polyangiitis (Churg-Strauss syndrome) as a special presentation of chronic rhinosinusitis with nasal polyps (CRSwNP). Allergol Select. 2024;8:18–25. doi: 10.5414/alx02475e.

5. Drynda A, Padjas A, Wójcik K, et al. Clinical characteristics of EGPA patients in comparison to GPA subgroup with increased blood eosinophilia from POLVAS registry. J Immunol Res. 2024;2024:4283928. doi: 10.1155/2024/4283928.

6. Reggiani F, L’Imperio V, Calatroni M, Pagni F, Sinico RA. Renal involvement in eosinophilic granulomatosis with polyangiitis. Front Med (Lausanne). 2023;10:1244651. doi: 10.3389/fmed.2023.1244651.

7. Comarmond C, Pagnoux C, Khellaf M, et al. Eosinophilic granulomatosis with polyangiitis (Churg-Strauss): clinical characteristics and long-term followup of the 383 patients enrolled in the French Vasculitis Study Group cohort. Arthritis Rheum. 2013;65(1):270–281. doi: 10.1002/art.37721.

8. Emmi G, Bettiol A, Gelain E, et al. Evidence-based guideline for the diagnosis and management of eosinophilic granulomatosis with polyangiitis. Nat Rev Rheumatol. 2023;19(6):378–393. doi: 10.1038/s41584-023-00958-w.

9. Watanabe R, Hashimoto M. Eosinophilic granulomatosis with polyangiitis: latest findings and updated treatment recommendations. J Clin Med. 2023;12(18):5996. doi: 10.3390/jcm12185996.

10. Vivek V, Yadav S, Korsapati HR, et al. Coronary artery dissection and myocarditis caused by eosinophilic granulomatosis with polyangiitis (EGPA): a case report. J Community Hosp Intern Med Perspect. 2023;13(5):50–56. doi: 10.55729/2000-9666.1219.

11. White JPE, Dubey S. Eosinophilic granulomatosis with polyangiitis: a review. Autoimmunity Rev. 2023;22(1):103219. doi: 10.1016/j.autrev.2022.103219.

12. Chung SA, Langford CA, Maz M, et al. 2021 American College of Rheumatology/Vasculitis Foundation guideline for the management of antineutrophil cytoplasmic antibody-associated vasculitis. Arthritis Rheumatol. 2021;73(8):1366–1383. doi: 10.1002/art.41773.

13. Guillevin L, Pagnoux C, Seror R, Mahr A, Mouthon L, Toumelin PL. The Five-Factor Score revisited: assessment of prognoses of systemic necrotizing vasculitides based on the French Vasculitis Study Group (FVSG) cohort. Medicine (Baltimore). 2011;90(1):19–27. doi: 10.1097/MD.0b013e318205a4c6.

14. Rice JB, White AG, Scarpati LM, Wan G, Nelson WW. Long-term systemic corticosteroid exposure: a systematic literature review. Clin Ther. 2017;39(11):2216–2229. doi: 10.1016/j.clinthera.2017.09.011.

15. Wechsler ME, Nair P, Terrier B, et al. Benralizumab versus mepolizumab for eosinophilic granulomatosis with polyangiitis. N Engl J Med. 2024;390(10):911–921. doi: 10.1056/NEJMoa2311155.

16. Terrier B, Jayne DRW, Hellmich B, et al. Clinical benefit of mepolizumab in eosinophilic granulomatosis with polyangiitis for patients with and without a vasculitic phenotype. ACR Open Rheumatol. 2023;5(7):354–363. doi: 10.1002/acr2.11571.

17. Jayne DRW, Terrier B, Hellmich B, et al. Mepolizumab has clinical benefits including oral corticosteroid sparing irrespective of baseline EGPA characteristics. ERJ Open Res. 2024;10(1):00509–02023. doi: 10.1183/23120541.00509-2023.

18. Hellmich B, Sanchez-Alamo B, Schirmer JH, et al. EULAR recommendations for the management of ANCA-associated vasculitis: 2022 update. Ann Rheum Dis. 2024;83(1):30–47. doi: 10.1136/ard-2022-223764.

19. Durel CA, Berthiller J, Caboni S, Jayne D, Ninet J, Hot A. Long-term followup of a multicenter cohort of 101 patients with eosinophilic granulomatosis with polyangiitis (Churg-Strauss). Arthritis Care Res (Hoboken). 2016;68(3):374–387. doi: 10.1002/acr.22686.

20. Bell CF, Blauer-Peterson C, Mao J. Burden of illness and costs associated with eosinophilic granulomatosis with polyangiitis: evidence from a managed care database in the United States. J Manag Care Spec Pharm. 2021;27(9):1249–1259. doi: 10.18553/jmcp.2021.21002.

21. Jakes RW, Kwon N, Nordstrom B, et al. Burden of illness associated with eosinophilic granulomatosis with polyangiitis: a systematic literature review and meta-analysis. Clin Rheumatol. 2021;40(12):4829–4836. doi: 10.1007/s10067-021-05783-8.

22. Gokhale M, Bell CF, Doyle S, Fairburn-Beech J, Steinfeld J, Van Dyke MK. Prevalence of eosinophilic granulomatosis with polyangiitis and associated health care utilization among patients with concomitant asthma in US commercial claims database. J Clin Rheumatol. 2021;27(3):107–113. doi: 10.1097/rhu.0000000000001198.

23. Hwee J, Harper L, Fu Q, Nirantharakumar K, Mu G, Jakes RW. Prevalence, incidence and healthcare burden of eosinophilic granulomatosis with polyangiitis in the UK. ERJ Open Res. 2024;10(3):00430–02023. doi: 10.1183/23120541.00430-2023.

24. Xu X, Edmonds C, Kim Y, et al. Disease overlap, healthcare resource utilization, and costs in patients with eosinophilic granulomatosis with polyangiitis: a REVEAL sub-study. Rheumatol Ther. 2024;11(6):1611–1628. doi: 10.1007/s40744-024-00714-w.

25. Ikeda M, Ohshima N, Kawashima M, Shiina M, Kitani M, Suzukawa M. Severe asthma where eosinophilic granulomatosis with polyangiitis became apparent after the discontinuation of dupilumab. Intern Med. 2022;61(5):755–759. doi: 10.2169/internalmedicine.7990-21.

26. US Department of Health and Human Services. Summary of the HIPAA Privacy Rule; 2013. Available from: https://www.hhs.gov/hipaa/for-professionals/privacy/laws-regulations/index.html. Accessed November 8, 2023.

27. Reddel HK, Bacharier LB, Bateman ED, et al. Global Initiative for Asthma Strategy 2021: Executive Summary and Rationale for Key Changes. J Allergy Clin Immunol Pract. 2022;10(1s):S1–S18. doi: 10.1016/j.jaip.2021.10.001.

28. Wechsler ME, Akuthota P, Jayne D, et al. Mepolizumab or Placebo for Eosinophilic Granulomatosis with Polyangiitis. N Engl J Med. 2017;376(20):1921–1932. doi: 10.1056/NEJMoa1702079.

29. Zou G. A modified poisson regression approach to prospective studies with binary data. Am J Epidemiol. 2004;159(7):702–706. doi: 10.1093/aje/kwh090.

30. Frome EL, Checkoway H. Epidemiologic programs for computers and calculators. Use of Poisson regression models in estimating incidence rates and ratios. Am J Epidemiol. 1985;121(2):309–323. doi: 10.1093/oxfordjournals.aje.a114001.

31. Tan L, Reibman J, Ambrose C, et al. Clinical and economic burden of uncontrolled severe noneosinophilic asthma. Am J Manag Care. 2022;28(6):e212–e220. doi: 10.37765/ajmc.2022.89159.

32. Yates M, Watts RA, Bajema IM, et al. EULAR/ERA-EDTA recommendations for the management of ANCA-associated vasculitis. Ann Rheum Dis. 2016;75(9):1583–1594. doi: 10.1136/annrheumdis-2016-209133.

