## Supplementary Materials for "Healthcare Resource Utilization and Costs for Patients With Eosinophilic Granulomatosis With Polyangiitis in the United States: A Retrospective Analysis of Health Insurance Claims Data"

### Supplementary Methods

#### Incident EGPA case definition

Persons with at least one inpatient diagnosis or two outpatient diagnoses (≥90 days apart) of eosinophilic granulomatosis with polyangiitis (EGPA) (International Classification of Diseases, Tenth Revision, Clinical Modification [ICD-10-CM] code M30.1) during the study selection window (January 1, 2017, through June 30, 2021) in the Merative™ MarketScan® Commercial Claims and Encounters database or Medicare Supplemental databases. Patients were excluded who had the EGPA diagnosis prior to January 1, 2017 or a first observed diagnosis of microscopic polyarteritis nodosa (ICD-10-CM code M31.7), giant cell arteritis (ICD-10-CM codes M31.5 and M31.6), Takayasu arteritis (ICD-10-CM code M31.4), or hypereosinophilic syndrome (ICD-10-CM D72.11) after the last observed EGPA diagnosis.

#### Matched severe uncontrolled asthma cohort definition

<u>Inclusion Criteria</u>

Patients were available for selection if they met all of the following criteria on or after January 1, 2017:

- At least one criterion from Part A, plus at least one criterion from Part B, plus at least one criterion from Part C (note: the criteria for Parts B and C must have occurred on or after January 1, 2017; the criteria from Part C must have occurred after the patient fulfilled the criteria from Part B. The criteria from Part A may have occurred at any point in time, including before January 1, 2017). The index date (ID) was determined based on Part C criteria, as described following.

**Part A:**

- At least one claim for an emergency department visit with a diagnosis of asthma (ICD-10-CM code J45.), or

- At least one claim for hospitalization with asthma as the primary diagnosis on the claim, or

- At least four outpatient claims on different dates (at least 7 days apart) with an asthma diagnosis in any position on the claim plus at least two pharmacy claims for asthma controller medications (inhaled corticosteroid [ICS], ICS in combination with a long-acting $\beta_2$ agonist [LABA], leukotriene receptor antagonist [LTRA] or theophylline) in 1 year (note: the four outpatient claims and two pharmacy claims must have all occurred within 365 days of each other; claims may have occurred in any order; outpatient claims must have been at least 7 days apart from one another; pharmacy claims must have been at least 60 days apart from each other, and no minimum number of days' supply was required for pharmacy claims), or

- At least one outpatient claim with an asthma diagnosis in any position, plus at least four pharmacy claims for any asthma medication in 1 year (note: the one outpatient claim and four pharmacy claims must all have occurred within 365 days of each other; claims may have occurred in any order; pharmacy claims must have been at least 60 days apart from one another, and no minimum number of days' supply was required for pharmacy claims).

**Part B:**

- Receipt of at least one claim with a medium- to high-dose of ICS, ICS/LABA, ICS/long-acting muscarinic antagonist (LAMA), or ICS/LTRA.

- At least 180 cumulative days' supply of oral glucocorticoids (OGCs); gaps present between fills that totaled ≥180 days were allowed.

**Part C:**

- At least one claim for hospitalization with asthma as the primary diagnosis on the claim (note: this could have been the same hospitalization as identified for Part A, so long as it occurred after the Part B criteria; the ID was defined as the date of admission), or

- At least two outpatient visits (ie, excluding inpatient stays) for asthma followed by treatment with systemic corticosteroids within a week (1 dose of intravenous steroids or at least 3 days of OGCs) (note: outpatient visits should have occurred between 30 and 365 days apart, provided that they occurred after the Part B criteria were fulfilled; each outpatient visit should have steroids dispensed within 1 week of the visit; the ID was defined as the date of the first outpatient visit with steroids dispensed), or

- At least four claims for a short-acting β2 agonist (SABA) within any 12-month period (note: the four SABA claims within a 12-month period should have been identified after the Part B criteria were met; each pharmacy claim was at least 30 days apart, and the ID was defined as the date of the first SABA claim).

Index Date

The ID was defined as the date the first Part C criterion was fulfilled on or after January 1, 2017 (note: all patients must have fulfilled the Part B criteria before the Part C criteria could be assessed). Patients were required to have at least 1 year of continuous health plan enrolment before ID.

Exclusion Criteria

Patients were excluded from the matched asthma cohort if they had a diagnosis of EGPA (ICD-10-CM code M30.1) at any time.

**Supplementary Table 1** Comorbidity Diagnosis Codes

| Condition | ICD-10-CM Code |
|---|---|
| Acute myeloid leukemia | C92.0, C92.4, C92.5, C92.6, C92.A |
| Anxiety | F06.4, F40, F41, F43.1 |
| Arrhythmia | I47, I48, I49 |
| Arterial thrombosis | I63.0, I63.3, I74 |
| Asthma | J45 |
| Atopic dermatitis, eczema | L20, L30.9 |
| Back pain (dorsalgia) | M54.0, M54.1, M54.4, M54.5, M54.6, M54.8, M54.9 |
| B-cell lymphoma | C82, C83, C85, C88.0, C88.2, C88.3, C88.4 |
| Cervical cancer | C53 |
| Chronic eosinophilic pneumonia | J82.81 |
| Chronic myeloid leukemia | C92.1, C92.2 |
| Chronic obstructive pulmonary disease | J41, J42, J43, J44, J98.2 |
| Colorectal cancer | C18, C19, C20 |
| Deep vein thromboembolism | I80.1, I80.2, I80.3, I81, I82.2, I82.3, I82.4, I82.5, I82.62, I82.72, I82.9 |
| Depression | F06.31, F06.32, F25.1, F31.3, F31.4, F31.5, F31.6, F31.7, F31.8, F31.9, F32, F33, F34.1, F41.8, F43.21, F43.23 |
| Diffuse eosinophilic fasciitis | M35.4 |
| Dyslipidemia | E78, E88.81 |
| Eosinophilic asthma | J82.83 |
| Eosinophilic cellulitis | L98.3 |
| Eosinophilic colitis | K52.82 |
| Eosinophilic endomyocardial disease | I42.3 |
| Eosinophilic esophagitis | K20.0 |
| Eosinophilic gastritis or gastroenteritis | K52.81 |
| Gastro-oesophageal reflux disease | K21 |
| Hodgkin’s lymphoma | C81 |
| Hypereosinophilic syndrome | D72.11 |
| Hyperthyroidism | E05 |
| Insomnia | F51.0, G47.0 |
| Interstitial pulmonary disease | J84, D86.0, D86.2 |
| Irritable bowel syndrome | K58 |
| Ischemic heart disease | I20, I21, I22, I23, I24, I25 |

| | |
|---|---|
| Lung cancer | C34 |
| Malignancy (all cancers) | C |
| Nasal polyps | J33 |
| Obesity | E66.0, E66.1, E66.2, E66.8, E66.9, Z68.3, Z68.4 |
| Obstructive sleep apnea | G47.33 |
| Polycythemia | D45 |
| Primary myelofibrosis | D47.1 |
| Primary (essential) thrombocythemia | D47.3 |
| Pulmonary eosinophilia | J82 |
| T-cell lymphoma | C84, C86, C91.5 |
| Throat or chest pain | R07 |
| Vitamin D deficiency | E55 |

**Abbreviations:** ICD-10-CM, International Classification of Diseases, Tenth Revision, Clinical Modification.

**Supplementary Table 2** BVAS Events

| Organ System | Event (ICD-10-CM code) |
|---|---|
| Major BVAS events | Gastrointestinal ischemia or infarction (K55.0, K55.1, K55.8, K55.9) |
| | Retinal change (retinal vasculitis, thrombosis, exudate or hemorrhage) (H34, H35.0, H35.2, H35.6, H35.7) |
| | Sensorineural hearing loss (H90.3, H90.4, H90.5, H90.6, H90.7, H90.8, H90.A2, H90.A3) |
| | Hemoptysis or alveolar hemorrhage (R04.2, R04.89) |
| | Respiratory failure (J96) |
| | Ischemic cardiac pain (I20.9) |
| | Cardiomyopathy (I25.5, I42.0, I42.1, I42.2, I42.3, I42.4, I42.5, I42.7, I42.8, I42.9, I43, O90.3) |
| | Congestive heart failure (I11.0, I13.0, I13.2, I26.0, I27.22, I50) |
| | Hematuria (N02, R31) |
| | Creatinine: stage IV–V chronic kidney disease, end-stage renal disease, or dialysis (G0049, G0052, G0257, G2170, G2171) |
| | Cerebrovascular accident (I60, I61, I62, I63, I67.6, I68, I69) |
| | Spinal cord lesion (G95.1) |
| | Cranial nerve palsy (G51.0, G51.2, H49.0, H49.1, H49.2, H49.3) |
| | Mononeuritis multiplex (G58.7) |
| Non-major BVAS events | |
| General | Myalgia (M79.1)<br>Arthralgia (M25.5)<br>Fever (R50.81, R50.9) |
| Skin | Purpura (D69.0, D69.2)<br>Skin ulcer (L97, L98.4, E08.621, E08.622, E09.621, E09.622, E10.621, E10.622, E11.621, E11.622, E13.621, E13.622)<br>Urticaria (L50)<br>Nodules (R22)<br>Erythema nodosum (L52) |
| Mucous membrane | Mouth ulcers (K12.3)<br>Genital ulcers (N48.5, N76.5, N76.6) |
| Eyes | Other specified disorders of eye and adnexa (H57.89)<br>Proptosis (H05.2)<br>Scleritis or episcleritis (H15.0, H15.1)<br>Conjunctivitis, blepharitis or keratitis (H01.0, H10, H16)<br>Moderate/severe visual impairment or blindness (H54.0, H54.1, H54.2, H54.3, H54.7, H54.8) |

| | |
|---|---|
| | Uveitis (H20) |
| Ear, nose, and throat | Bloody nasal discharge, crusts, nasal ulcers or nasal granulomata (J34.0, J34.8)<br>Subglottic stenosis (J38.6)<br>Conductive hearing loss (H90.0, H90.1, H90.2, H90.6, H90.7, H90.8, H90.A1, H90.A3)<br>Acute or chronic sinusitis (J01, J32) |
| Chest | Wheeze (R06.2)<br>Lung nodules or cavities (R91)<br>Pleural effusion or pleurisy/pleuritis (J90, J91.8, R09.1)<br>Endobronchial involvement (J98.09) |
| Cardiovascular | Valvular heart disease (I34, I35, I36, I37)<br>Pericarditis or pericardiectomy (I30.0, I30.8, I30.9, I31.0, I31.1, I31.9) |
| Abdominal | Peritonitis (K65) |
| Renal | Hypertension (I10, I11, I12, I13, I14, I16, I67.4)<br>Proteinuria (R80.0, R80.1, R80.9)<br>Creatinine: stage II–III chronic kidney disease (I12.9, I13.0, I13.10, N18.2, N18.3, N18.9) |
| Nervous system | Headache (G44.0, G44.1, G44.5, G44.8, R51)<br>Meningitis (G03)<br>Organic confusion (R41.0)<br>Seizure (G40, R56.9)<br>Sensory peripheral neuropathy (R20.2) |

**Note:** Events were based on version 3.0 of the BVAS.

**Abbreviation:** BVAS, Birmingham Vasculitis Activity Score.

**Supplementary Table 3** Demographics, Clinical Characteristics, and Insurance Type for the Full Incident EGPA Cohort (Prior to Match), Matched EGPA Cohort A, and Matched General Insured Cohort

| | **Full Incident EGPA Cohort (N = 236)** | **Matched EGPA Cohort A (n = 213)** | **Matched General Insured Cohort (n = 779)** |
|---|---|---|---|
| Age at index date, mean (SD) | 50.4 (14.5) | 49.6 (14.3) | 50.6 (13.7) |
| [range], years | [1.0–85.0] | [1.0–85.0] | [2.0–85.0] |
| Age at index date, n (%) | | | |
| <18 years | 4 (1.7) | 4 (1.9) | 12 (1.5) |
| 18–34 years | 34 (14.4) | 31 (14.6) | 78 (10.0) |
| 35–49 years | 66 (28.0) | 66 (31.0) | 242 (31.1) |
| 50–64 years | 104 (44.1) | 90 (42.3) | 372 (47.8) |
| 65–79 years | 23 (9.7) | 18 (8.5) | 62 (8.0) |
| 80+ years | 5 (2.1) | 4 (1.9) | 13 (1.7) |
| Female, n (%) | 136 (57.6) | 125 (58.7) | 459 (58.9) |
| Follow-up duration from index date, mean (SD), months | 21.7 (14.6) | 20.8 (14.3) | 20.7 (14.3) |
| Duration of follow-up from index date, n (%) | | | |
| <6 months | 39 (16.5) | 38 (17.8) | 145 (18.6) |
| 6–11 months | 39 (16.5) | 37 (17.4) | 128 (16.4) |
| 12–17 months | 31 (13.1) | 28 (13.1) | 107 (13.7) |
| 18–23 months | 33 (14.0) | 30 (14.1) | 113 (14.5) |
| 24–29 months | 32 (13.6) | 30 (14.1) | 110 (14.1) |
| 30+ months | 62 (26.3) | 50 (23.5) | 176 (22.6) |
| Year of index date, n (%) | | | |
| 2017 | 67 (28.4) | 59 (27.7) | 223 (28.6) |
| 2018 | 50 (21.2) | 44 (20.7) | 158 (20.3) |
| 2019 | 63 (26.7) | 58 (27.2) | 210 (27.0) |
| 2020 | 37 (15.7) | 34 (16.0) | 120 (15.4) |
| 2021 | 19 (8.1) | 18 (8.5) | 68 (8.7) |
| Commercial insurance, n (%) | 208 (88.1) | 191 (89.7) | 703 (90.2) |
| Medicare Supplement/Medicare Advantage, n (%) | 28 (11.9) | 22 (10.3) | 76 (9.8) |
| Geographic region, n (%) | | | |
| Northeast | 50 (21.2) | 43 (20.2) | 156 (20.0) |
| North Central | 42 (17.8) | 41 (19.2) | 155 (19.9) |
| South | 97 (41.1) | 86 (40.4) | 317 (40.7) |
| West | 47 (19.9) | 43 (20.2) | 151 (19.4) |

**Abbreviations:** EGPA, eosinophilic granulomatosis with polyangiitis; SD, standard deviation.

**Supplementary Table 4** History of Comorbidities in Matched EGPA Cohort A and the Matched General Insured Cohort

| Comorbidity | Percentage Prevalence (95% CI) | | Prevalence Ratio (95% CI) | *P*-value |
|---|---|---|---|---|
| | Matched EGPA Cohort A (n = 213) | Matched General Insured Cohort (n = 779) | | |
| Asthma | | | | |
| Persistent | 70.4 (64.6, 76.8) | 2.7 (1.8, 4.1) | 26.1 (17.0, 40.2) | < .001 |
| Severe | 65.7 (59.7, 72.4) | 1.2 (0.6, 2.2) | 56.9 (29.5, 109.7) | < .001 |
| Severe uncontrolled | 45.1 (38.9, 52.3) | 0.4 (0.1, 1.2) | 117.0 (37.5, 365.6) | < .001 |
| Throat or chest pain | 46.0 (39.8, 53.2) | 17.1 (14.6, 19.9) | 2.7 (2.2, 3.3) | < .001 |
| Dyslipidemia | 40.4 (34.3, 47.5) | 39.2 (35.9, 42.7) | 1.0 (0.9, 1.2) | .745 |
| Gastro-oesophageal reflux disease | 37.1 (31.1, 44.2) | 16.7 (14.3, 19.5) | 2.2 (1.8, 2.8) | < .001 |
| Back pain | 32.9 (27.1, 39.8) | 26.4 (23.5, 29.7) | 1.2 (1.0, 1.6) | .058 |
| Nasal polyps | 30.0 (24.5, 36.9) | 0.5 (0.2, 1.4) | 58.5 (21.6, 158.9) | < .001 |
| Vitamin D deficiency | 25.4 (20.1, 31.9) | 17.6 (15.1, 20.5) | 1.4 (1.1, 1.9) | .009 |
| Obesity | 25.4 (20.1, 31.9) | 23.6 (20.8, 26.8) | 1.1 (0.8, 1.4) | .598 |
| Anxiety | 25.4 (20.1, 31.9) | 20.9 (18.3, 24.0) | 1.2 (0.9, 1.6) | .160 |
| COPD | 19.7 (15.0, 25.9) | 3.0 (2.0, 4.4) | 6.7 (4.1, 10.9) | < .001 |
| Arrhythmia | 19.7 (15.0, 25.9) | 6.5 (5.0, 8.5) | 3.0 (2.1, 4.4) | < .001 |
| Obstructive sleep apnea | 19.7 (15.0, 25.9) | 14.8 (12.5, 17.5) | 1.3 (1.0, 1.8) | .076 |

| | | | | |
|---|---|---|---|---|
| Atopic dermatitis/eczema | 18.3 (13.8, 24.3) | 6.5 (5.0, 8.5) | 2.8 (1.9, 4.1) | < .001 |
| Depression | 17.8 (13.4, 23.8) | 16.4 (14.0, 19.3) | 1.1 (0.8, 1.5) | .624 |
| Ischemic heart disease | 17.8 (13.4, 23.8) | 5.4 (4.0, 7.2) | 3.3 (2.2, 5.0) | < .001 |
| Pulmonary eosinophilia | 16.9 (12.5, 22.8) | 0.1 (0.0, 0.9) | 131.7 (18.2, 954.7) | < .001 |
| Interstitial pulmonary disease | 12.2 (8.5, 17.5) | 0.3 (0.1, 1.0) | 47.5 (11.4, 198.7) | < .001 |
| Any malignancy | 10.3 (7.0, 15.3) | 6.8 (5.2, 8.8) | 1.5 (0.9, 2.4) | .084 |
| Deep vein thromboembolism | 5.2 (2.9, 9.2) | 0.9 (0.4, 1.9) | 5.7 (2.3, 14.6) | < .001 |
| Primary (essential) thrombocythemia | 3.8 (1.9, 7.4) | 0.3 (0.1, 1.0) | 14.6 (3.1, 68.4) | < .001 |
| Eosinophilic esophagitis | 3.8 (1.2, 6.3) | 0 | – | – |
| Irritable bowel syndrome | 3.3 (1.6, 6.8) | 3.3 (2.3, 4.9) | 1.0 (0.4, 2.2) | .971 |
| Hyperthyroidism | 3.3 (1.6, 6.8) | 0.4 (0.1, 1.2) | 8.5 (2.2, 32.7) | .002 |
| Hypereosinophilic syndrome | 2.3 (0.3, 4.4) | 0 | – | – |
| Eosinophilic gastritis or gastroenteritis | 1.4 (0.0, 3.0) | 0 | – | – |
| Eosinophilic endomyocardial disease | 1.4 (0.0, 3.0) | 0 | – | – |

**Note:** History of comorbidities in the baseline period (12 months prior to the index date) occurring in ≥ 1% of patients in either Matched EGPA Cohort A or the Matched General Insured Cohort is shown.
**Abbreviations:** CI, confidence interval; COPD, chronic obstructive pulmonary disease; EGPA, eosinophilic granulomatosis with polyangiitis.

**Supplementary Table 5** Demographics, Clinical Characteristics, and Insurance Type for the Full Incident EGPA Cohort (Prior to Match), Matched EGPA Cohort B, and Matched SUA Cohort

| | Full Incident EGPA Cohort (N = 236) | Matched EGPA Cohort B (n = 182) | Matched SUA Cohort (n = 640) |
|---|---|---|---|
| Age at index date, mean (SD) [range], years | 50.4 (14.5) [1.0–85.0] | 51.2 (14.0) [1.0–85.0] | 51.7 (13.6) [2.0–86.0] |
| Age at index date, n (%) | | | |
| <18 years | 4 (1.7) | 4 (2.2) | 11 (1.7) |
| 18–34 years | 34 (14.4) | 22 (12.1) | 66 (10.3) |
| 35–49 years | 66 (28.0) | 50 (27.5) | 168 (26.3) |
| 50–64 years | 104 (44.1) | 86 (47.3) | 325 (50.8) |
| 65–79 years | 23 (9.7) | 15 (8.2) | 55 (8.6) |
| 80+ years | 5 (2.1) | 5 (2.7) | 15 (2.3) |
| Female, n (%) | 136 (57.6) | 108 (59.3) | 390 (60.9) |
| Follow-up duration from index date, mean (SD), months | 21.7 (14.6) | 19.9 (13.3) | 18.8 (12.6) |
| Duration of follow-up from index date, n (%) | | | |
| <6 months | 39 (16.5) | 31 (17.0) | 111 (17.3) |
| 6–11 months | 39 (16.5) | 33 (18.1) | 132 (20.6) |
| 12–17 months | 31 (13.1) | 27 (14.8) | 99 (15.5) |
| 18–23 months | 33 (14.0) | 29 (15.9) | 104 (16.3) |
| 24–29 months | 32 (13.6) | 24 (13.2) | 76 (11.9) |
| 30+ months | 62 (26.3) | 38 (20.9) | 118 (18.4) |
| Year of index date, n (%) | | | |
| 2017 | 67 (28.4) | 49 (26.9) | 171 (26.7) |
| 2018 | 50 (21.2) | 42 (23.1) | 154 (24.1) |
| 2019 | 63 (26.7) | 50 (27.5) | 168 (26.3) |
| 2020 | 37 (15.7) | 29 (15.9) | 106 (16.6) |
| 2021 | 19 (8.1) | 12 (6.6) | 41 (6.4) |
| Commercial insurance, n (%) | 208 (88.1) | 162 (89.0) | 569 (88.9) |
| Medicare Supplement/Medicare Advantage, n (%) | 28 (11.9) | 20 (11.0) | 71 (11.1) |
| Geographic region, n (%) | | | |
| Northeast | 50 (21.2) | 40 (22.0) | 150 (23.4) |
| North Central | 42 (17.8) | 33 (18.1) | 123 (19.2) |
| South | 97 (41.1) | 74 (40.7) | 249 (38.9) |
| West | 47 (19.9) | 35 (19.2) | 118 (18.4) |

**Abbreviations:** EGPA, eosinophilic granulomatosis with polyangiitis; SD, standard deviation; SUA, severe uncontrolled asthma.

**Supplementary Table 6** History of Comorbidities in the Matched EGPA Cohort B and Matched SUA Cohort

| Comorbidity | Percentage Prevalence (95% CI) | | Prevalence Ratio (95% CI) | *P*-value |
|---|---|---|---|---|
| | Matched EGPA Cohort B (n = 182) | Matched SUA Cohort[a] (n = 640) | | |
| Asthma | | | | |
| Persistent | 70.3 (63.7, 77.0) | – | – | – |
| Severe | 64.3 (57.3, 71.2) | – | – | – |
| Severe uncontrolled | 42.9 (35.7, 50.0) | – | – | – |
| Dyslipidemia | 43.4 (36.8, 51.2) | 48.9 (45.2, 52.9) | 0.9 (0.7, 1.1) | .203 |
| Throat or chest pain | 42.9 (36.2, 50.7) | 37.0 (33.5, 41.0) | 1.2 (1.0, 1.4) | .144 |
| Gastro-oesophageal reflux disease | 39.0 (32.5, 46.8) | 36.3 (32.7, 40.2) | 1.1 (0.9, 1.3) | .491 |
| Back pain | 31.9 (25.8, 39.4) | 41.1 (37.5, 45.1) | 0.8 (0.6, 1.0) | .032 |
| Nasal polyps | 28.0 (22.2, 35.4) | 4.7 (3.3, 6.6) | 6.0 (3.9, 9.1) | < .001 |
| Anxiety | 27.5 (21.7, 34.8) | 28.9 (25.6, 32.6) | 1.0 (0.7, 1.2) | .707 |
| Vitamin D deficiency | 26.4 (20.7, 33.6) | 23.3 (20.2, 26.8) | 1.1 (0.9, 1.1) | .384 |
| Obesity | 24.7 (19.2, 31.9) | 41.7 (38.1, 45.7) | 0.6 (0.5, 0.8) | < .001 |
| COPD | 21.4 (16.2, 28.3) | 22.7 (19.6, 26.1) | 0.9 (0.7, 1.3) | .727 |
| Depression | 20.3 (15.2, 27.1) | 23.3 (20.2, 26.8) | 0.9 (0.6, 1.2) | .407 |
| Arrhythmia | 19.8 (14.8, 26.5) | 10.5 (8.3, 13.1) | 1.9 (1.3, 2.7) | < .001 |
| Obstructive sleep apnea | 19.2 (14.3, 25.9) | 29.7 (26.4, 33.4) | 0.6 (0.5, 0.9) | .008 |

| | | | | |
|---|---|---|---|---|
| Ischemic heart disease | 18.7 (13.8, 25.3) | 12.0 (9.8, 14.8) | 1.6 (1.1, 2.2) | .019 |
| Atopic dermatitis/eczema | 18.7 (13.8, 25.3) | 11.9 (9.6 ,14.7) | 1.6 (1.1, 2.3) | .016 |
| Pulmonary eosinophilia | 16.5 (11.9, 22.9) | 0.5 (0.2, 1.4) | 35.2 (10.9, 113.9) | < .001 |
| Interstitial pulmonary disease | 12.6 (8.6, 18.5) | 3.1 (2.0, 4.8) | 4.0 (2.3, 7.2) | < .001 |
| Any malignancy | 11.0 (7.3, 16.6) | 8.8 (6.8, 11.2) | 1.3 (0.8, 2.0) | .355 |
| Deep vein thromboembolism | 4.9 (2.6, 9.3) | 2.3 (1.4, 3.9) | 2.1 (0.9, 4.7) | .071 |
| Hyperthyroidism | 4.9 (2.6, 9.3) | 1.6 (0.8, 2.9) | 3.2 (1.3, 7.7) | .011 |
| Primary (essential) thrombocythemia | 4.4 (2.2, 8.7) | 0.2 (0.0, 1.1) | 28.1 (3.5, 223.5) | .002 |
| Eosinophilic esophagitis | 3.8 (1.9, 8.0) | 1.1 (0.5, 2.3) | 3.5 (1.2, 9.9) | .017 |
| Irritable bowel syndrome | 3.3 (1.5, 7.2) | 5.5 (4.0, 7.5) | 0.6 (0.3, 1.4) | .243 |
| Eosinophilic endomyocardial disease | 2.2 (0.1, 4.3) | 0 | – | – |
| Arterial thrombosis | 1.6 (0.5, 5.1) | 0.5 (0.2, 1.4) | 3.5 (0.7, 17.3) | .122 |
| Eosinophilic gastritis or gastroenteritis | 1.6 (0.5, 5.1) | 0.2 (0.0, 1.1) | 10.5 (1.1, 100.8) | .041 |
| B-cell lymphoma | 1.1 (0.3, 4.4) | 0.8 (0.3, 1.9) | 1.4 (0.3, 7.2) | .0682 |
| Hypereosinophilic syndrome | 1.1 (0.0, 2.6) | 0 | – | – |

**Notes:** [a]All patients in the SUA cohort had persistent, severe, and severe uncontrolled asthma.
History of comorbidities in the baseline period (12 months prior to the index date) occurring in ≥ 1% of patients in either the EGPA Matched Cohort B or Matched SUA Cohort is shown.
**Abbreviations:** CI, confidence interval, COPD, chronic obstructive pulmonary disease; EGPA, eosinophilic granulomatosis with polyangiitis, SUA, severe uncontrolled asthma.

**Supplementary Figure 1 Study Design.**

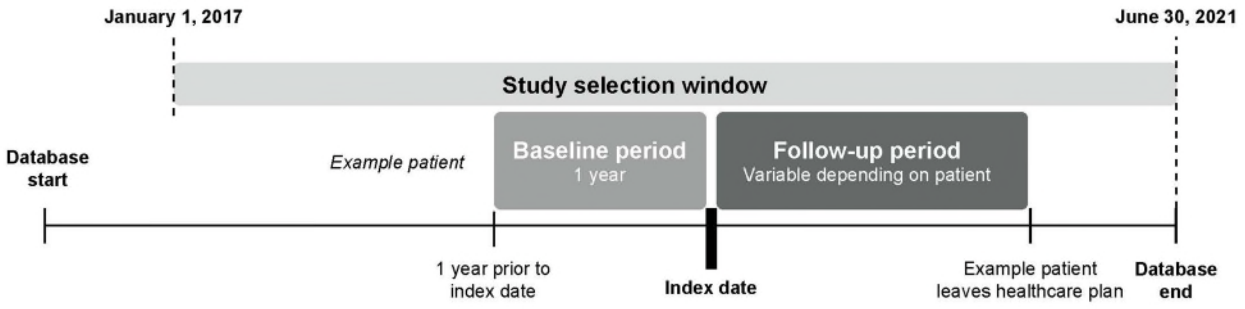

**Supplementary Figure 2** Patient selection and attrition for the Full Incident EPGA Cohort.

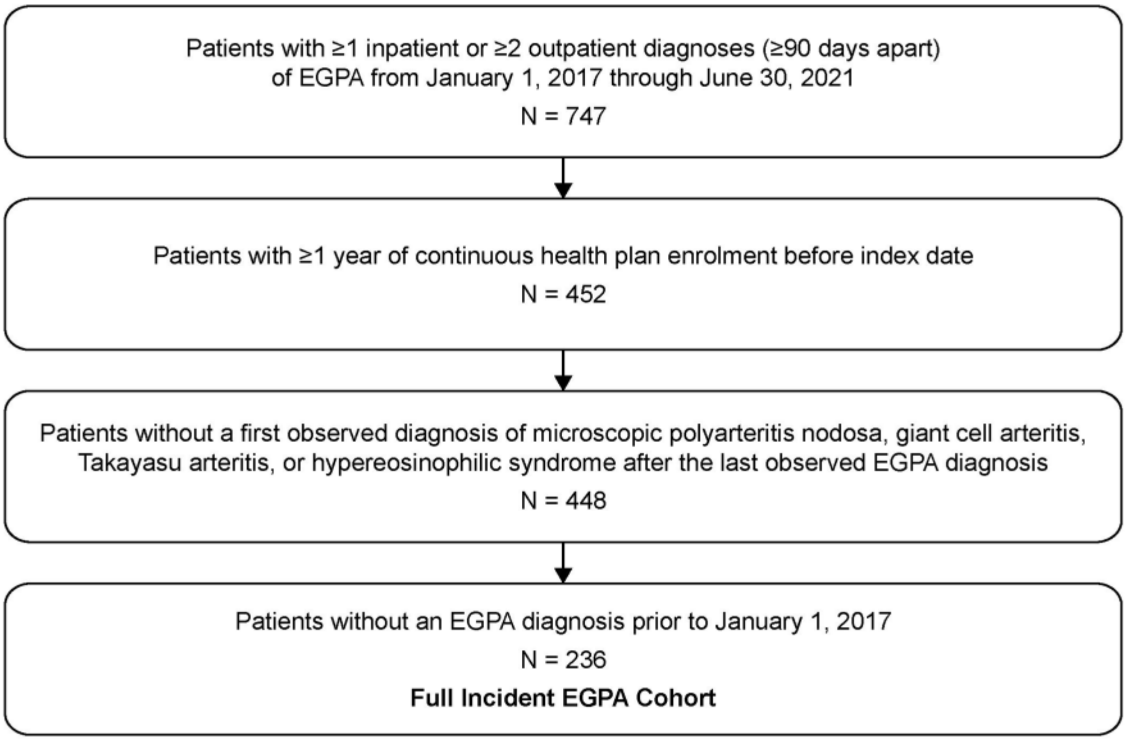


**Abbreviation:** EGPA, eosinophilic granulomatosis with polyangiitis.
